# The influence of skin pigmentation on pulse oximetry readings: a protocol for systematic review and meta-analysis

**DOI:** 10.1101/2023.10.18.23297133

**Authors:** Shabbir Ahmed, Sadia Afrin, SK Mehedi Hasan, Sharmishtha Sonalika Sarker, Farhia Azrin, Esrat Jahan, Ahmed Ehsanur Rahman

## Abstract

**Background:** Pulse oximeters are widely used crucial devices to triage and monitor sign of diseases and make informed decisions on patients’ health. Various studies have found a bias in pulse oximetry readings based on skin pigmentation/ ethnicity. This protocol outlines a systematic review and meta-analysis approach that will assess the effect of different skin pigmentation levels on the accuracy of pulse oximetry readings.

**Methods and analysis:** We will conduct search in Medline, Embase, CINAHL, and Web of Science database. Studies carried out either on healthy population or population with critical illness measuring pulse oximetry (SpO2) versus SaO2 obtained by standard CO-oximetry based on skin pigmentation and/or ethnicity and reported in the form of overall accuracy, bias, precision, or agreement will be included in this study. Four dedicated reviewers will independently assess and extract data from the included studies in a pre-specified excel format for further analysis. Any discordance in the way will be resolved by discussion until consensus is reached. JBI’s critical appraisal tool for diagnostic test accuracy test will be used to assess study quality for the included studies.

**Ethics and dissemination:** Ethical approval is not required because no original data will be collected as part of this review. The final review will be submitted to a peer-reviewed journal for publication and presented at conferences.

## Background

### Oxygen saturation measurement

Urgency to monitor blood oxygen saturation levels, in a multitude of circumstances, for health reasons is crucial. Precisely, low blood oxygen levels, known as hypoxemia, necessitates urgent medical attention and is associated with a higher risk of mortality (Swigris et al., 2009). Currently, CO-oximetry is the gold standard procedure to measure blood oxygen saturation level (SaO_2_). Additionally, the use of pulse oximetry, as a proxy measure for (SaO_2_), as a non-invasive and simple use of devices also in practice to measure blood oxygen levels known as SpO_2_. The wide introduction of pulse oximetry both in clinical and non-clinical settings in recent times has gained a tremendous acceleration and was highly in demand during the COVID-19 pandemic (NHS, 2023). Being non-invasive and relatively inexpensive has made it more in use as a proxy measure for (SaO_2_).

The basic mechanism of pulse oximeter bridged on the principle of the Beer-Lambert principle that dictates the attenuation of light to the properties of the material through which the light is travelling (Beer & Beer, 1852) (Chan, 2013). In the description of (Aoyagi, 2003) and (Sinex, 1999) pulse oximeter detects oxygen saturation by measuring transdermal light absorption in blood through skin, for example through the fingertip. The visible red part of the spectrum is absorbed by the reduced hemoglobin fraction and infrared fraction of the spectrum is absorbed by oxyhemoglobin fraction. Thus the relative proportional presence of these two fraction is calculated to produce the SpO2 value. There are two kinds of pulse oximetry are currently in use one is *transmissive*, where light emitter and detector of the pulse oximeter face each other, and the other is *reflectance*, where probe employs an emitter adjacent to the detector and rely on the signal being reflected or backscattered through the tissue.

### Brief orientation of skin pigmentation and ethnicity with pulse oximetry

Skin pigmentation primarily indicates the skin color or skin tone. Scientific classification like Fitzpatrick scale classifies skin pigmentation into six groups-light, pale white; white, fair; medium white to light brown; olive, moderate brown; brown, dark brown; and very dark brown (Fitzpatrick, 1988). Scientific communities are more consensus on the issue of rising concern regarding the performance of the pulse oximetry particularly in patients with different skin pigmentation (Shi et al., 2022). Though the concept of pulse oximetry accuracy based on different skin tone is not a relatively new discovery rather the urgency of this biased reading was deeply realized during the COVID-19 (Greenhalgh et al., 2021). Moreover, mortality burden during the COVID-19 among the ethnic minority patients with skin pigmentation has rightfully raised the question of pulse oximetry accuracy (Al-Halawani, 2023).

### Current evidence in clinical milieu

In order to triage and monitor the bulk of patients during COVID-19, deciding who needs the more critical care and therapy, pulse oximetry was a highly relied (Silverston, 2022). Due to the significant burden of COVID-19 illness and the danger of occult hypoxemia the convenient and low cost utility of the pulse oximetry made them more demanding (Luks & Swenson, 2020). Moreover, medical professionals dubbed pulse oximeter devices as a game changer and widely advised for the self-evaluation purpose (Levitan, 2020) (Mathur, 2020). Nonetheless, amid the preliminary clinical merit, there are suspicions relating their accuracy and potential shortcomings in interpreting the measured results (Luks & Swenson, 2020). For more than three decades there has been ongoing debates on the potential impact of darker skin pigmentation on accurate pulse oximetry reading. On top of that racial disparities in mortality and morbidity amid the pandemic have prompted questions about whether they might be, in part, caused by the difference in performance of pulse oximetry in patients with darker skin pigmentation (Silverston, 2022).

As mentioned earlier, over a span of 28 years ago, the first systematic review study pointed the issue of assessing the accuracy of pulse oximetry to measure oxygen saturation and the effect of factors including skin pigmentation on pulse oximetry. In this study among 74 included studies 44 studies reported agreement between pulse oximetry and the reference standard measurement while 27 studies outlined the bias and precision summarizing the finding that pulse oximeter were accurate within 2% (±1SD) or 5% (±2SD) of in vitro oximetry in the range of 70% to 100% SaO_2_. Thus the skin pigmentation effect on pulse oximetry readings were interpreted as skin with high level of skin pigmentation may cause over estimation of oxygen saturation with no evidence on ethnicity/race (Jensen, 1998). After that a recent study of Shi and colleagues (2022) further clarifies that compared with standard SaO_2_ measurement, pulse oximetry probably overestimates oxygen saturation in people with the high level of skin pigmentation (pooled mean bias 1.11%; 95% confidence interval 0.29 to 1.93%) and people described as Black/African American (1.52%; 0.95 to 2.09%) (moderate- and low-certainty evidence). The further confirmation of their findings dictates that the evidence of the measuring bias found for other skin pigmentation levels or races is still ambiguous (Shi et al., 2022) which further fuel for the clarity on the crucial aspect of overestimation/underestimation or variation in the readings of pulse oximetry.

In order to provide required medical attention for the critical patients with skin pigmentation, based on the discussion, it is necessary to ensure the inclusion of all relevant information available to date to make informed decision and further provocation of knowledge regarding the pulse oximetry readings. Thus this proposed review will assess the effect of different skin pigmentation levels on the accuracy of pulse oximetry readings.

### Review objective

To assess the effects of skin pigmentation and race/ethnicity, however defined, on the reliability of oxygen saturation measurement by pulse oximetry (SpO_2_) compared with SaO_2_ measure by standard CO-oximetry.

## Methods

To aid in the design of this targeted review strategy, we first carried out an informal scoping exercise utilizing the search and screening procedure described below. Any study that compared the SpO_2_ readings from various pulse oximeters with the SaO_2_ readings in any cohort was included in the scope.

### Criteria for considering studies for this review

#### Inclusion criteria

##### Study types

We will include all studies providing original observational data, including observational studies and interventional studies reporting overall accuracy, bias, precision, or agreement data on oxygen saturation assessed by pulse oximetry (SpO_2_) vs SaO_2_ obtained by standard CO-oximetry based on skin pigmentation and/or ethnicity.

##### Participant

Inclusion of any population with any level of baseline oxygen saturation treated in any care/clinical settings will be targeted with the specification of

- Population (children and adults) with multitudes of health complexities (for example patients with chronic illness and patients went under surgical operations
- Population (children and adults) without any health conditions (for example healthy volunteers and athletes)

##### Skin pigmentation definition

As the study is focused on finding the impact of skin pigmentation levels on pulse oximetry readings the inclusion of participants with mentioned criteria with the level of skin pigmentation reported in Fitzpatrick scale will be considered. Additionally, we will also include papers reporting any subjective measures/judgement of the skin pigmentation such as dark or light pigmentation. Moreover, we will also include skin pigmentation reported in Munsell scale and measured in portable EEL reflectance spectrometer. Besides, papers reporting race/ethnicity to indicate skin pigmentation or in itself will also be included in this study.

##### Eligible review comparison

Assessing the accuracy of the pulse oximeter based estimation of oxygen saturation (SpO_2_) compared with the gold standard measure of SaO_2_ poised by standard CO-oximetry.

##### Pulse oximetry devices

Studies that measure SpO_2_ value at any care or clinical settings with the use of following pulse oximeters for measuring oxygen saturation will be prioritized. Additionally, eligible oximeters included in this study can have either feature of reflectance probe or a transmissive probe.

- Standalone commercial pulse oximeter with the probe and monitor as a simple and single assembly such as fingertip, handheld or tabletop devices
- Reversely, pulse oximeter consisting of probe and monitor as separate or portable parts
- Monitoring devices with portability functions for either for home use or professional use with pulse oximetry test
- Wearable devices (e.g. smartwatch, wrist braces)
- Smart-phone mechanized devices with pulse oximetry test.

##### Comparator measurement

Blood oxygen saturation, as measured peripherally by pulse oximetry (SpO_2_) or arterially by blood gas analysis (SaO_2_). Thus we will include studies that uses the gold standard method of measuring arterial blood oxygen saturation (SaO_2_) blood gas analysis with CO-oximetry.

##### Eligible outcomes

Outcomes for this study will be measured based on the accumulated information of the following criteria

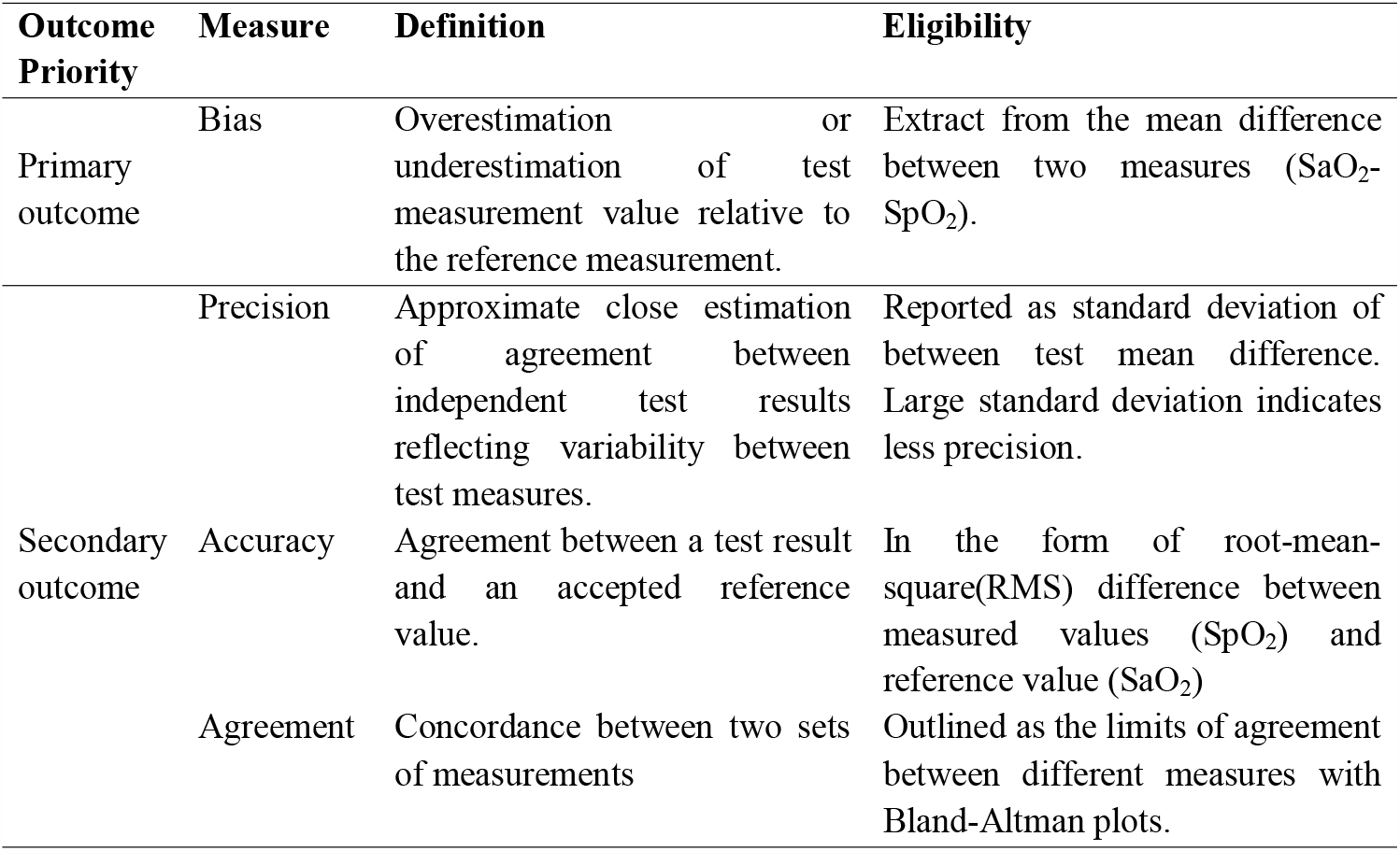

#### Exclusion criteria

- Studies that use pulse oximetry as a diagnostic tool to define hypoxia based on a predetermined oxygen saturation threshold in comparison to the gold standard value will be excluded.
- We will also exclude studies in which prototype pulse oximetry devices, pulse oximeters that require high skilled specialists to operate (such as intra-partum pulse oximetry devices), and pulse oximeters is used for measuring venous blood oxygen saturation.
- Furthermore, we will exclude measurements with venous blood samples, mixed arterial and venous such as capillary blood and earlobe blood and also arterial blood or umbilical arterial blood that requires highly skilled specialists/advanced equipment to sample.

More specifically we will exclude

- Blood gas analyzers that measures PaO_2_ (arterial oxygen pressure) for calculation of SaO_2_ values, rather than measuring SaO_2_ directly
- Other reference comparators values of oxygen saturation such as O_2_Hb or FO_2_Hb.
- Calibrated pulse oximetry readings

- We will exclude studies reported blood gas analyzers that measures PaO_2_ (arterial oxygen pressure) for calculation of SaO_2_ values, rather than measuring SaO_2_ directly. Other reference comparators values of oxygen saturation such as O_2_Hb or FO_2_Hb will also be excluded with additional attention on calibrated pulse oximetry readings
- Additionally, in seeking eligible outcomes studies will be excluded with reported estimation presented in regression and correlation measures. Moreover, studies on repeatability or reliability without comparison to a reference standard (SaO2) will not be included.

### Search databases and strategies for identification of studies

#### Search database

From 2022 onward, we will conduct searches and look through the following electronic database to find targeted studies

- Medline
- Embase
- CINAHL
- Web of Science

#### Database search

Boolean operators like ‘AND,’ and ‘OR,’ will be used in subject header and free text searches to correspond with the eligibility criteria for the systematic review. The search terms will include ‘Pulse oximeter OR oximetry OR SpO_2_’, AND ‘CO-oximetry OR blood measure OR gold standard OR blood analysis’, AND ‘bias/agreement OR comparative study OR sensitivity and specificity’.

Each database’s search method will be modified using specific search terms. Furthermore, we won’t restrict our search by language but rather by publishing year. We will contact the authors to retrieve any additional information about the studies if necessary. The Supplementary material contains a detailed search technique for each database, along with any relevant specifics.

### Study selection

#### Review of titles and abstract

Once duplication has been identified and eliminated four dedicated reviewers will independently assess the titles and abstract of the search results for relevance with a meticulous and rigorous attention of the eligible review comparator and outcome set for this review. If the eligibility criteria are met corresponding studies will be marked as ‘yes’ otherwise ‘no’ and will be followed for going through to full-text review in the next phase.

#### Full text review

At this point, the inclusion and exclusion criteria will be thoroughly examined. All articles that make it to this stage will be saved in a shared folder for the final review. If additional information is required, the authors of the articles will be contacted. When the screening is finished, a flow chart will be created using the PRISMA guidelines to summarize the screening. Additionally, minor disputes will be resolved by the process of cross-check among the reviewers interchangeably until consensus point is met.

### Data extraction

We will use Excel for the title and abstracts screening. After the initial title–abstract screening, full-text articles will be obtained of all potential studies for full text review. Upon the selection for the full-text review based on specified inclusion/exclusion criteria all the four reviewers will be independently performed the data extraction procedure manually with a structured set of variables in a printed format. A pilot will be conducted with one article to ensure the quality of the extraction. All the printed data extraction formats will further be compiled in spreadsheet for a comprehensive overview and for the convenience of analysis. Resolve of any disagreements will be reached through crosscheck and discussion.

Printed data extracted format for the selected studies will have variables including

- Basic characteristics of studies such author name, journal name, publication year, study type and study region
- Study settings (e.g. inpatient or outpatient, laboratory setting, emergency, community setting or any other)
- Population characteristics (e.g. age and sex composition, clinical conditions etc.)
- Skin pigmentation definitions including scales used and pigmentation levels as well as the race/ethnicity
- Oximeter characteristics, probe sites
- Methods for measuring SaO2 including the blood gas analyzer and CO-oximetry model and blood source
- Data on accuracy, bias, and precision of measurement at comparative level of skin pigmentation and race/ethnicity group
- Other factors reported to have the effects on pulse oximetry accuracy.

Additionally, necessary data transformation techniques will be applied for the convenient of analysis purpose (for example: from reported 95% confidence interval to standard deviation).

Clarification of any dispute in data extraction will be resolved through contacting authors and deciding for an inclusion and exclusion decision.

### Risk of bias assessment

JBI’s (Campbell, 2020) critical appraisal tool for diagnostic test accuracy test will be used to assess study quality.

### Data synthesis strategy

Primarily data will be entered into Microsoft Excel, and analysis will be done by Stata 17 (StataCorp, 2021). Summaries of the included studies and data narrative will further be synthesized using meta-analysis.

### Sub group analysis

Based on the findings of extracted data, additional outcomes will be analyzed as sub-groups or subsets to present country or location-specific analyses.

### Sensitivity analysis

In order to assess the robustness of the meta-analysis sensitivity analysis will be followed.

### Publication bias

We will assess publication bias using funnel plots and Egger’s tests (Sterne, 2001).

## Data Availability

All data produced in the present study are available upon reasonable request to the authors

## List of Abbreviations

COVID-19: Coronavirus disease 2019
SD: Standard deviation
PRISMA: Preferred Reporting Items for Systematic Reviews and Meta-Analyses
JBI: Joanna Biggs Institute

## Declarations

### Ethics approval and consent to participate

Not applicable to this protocol.

### Consent for publication

Not applicable to this protocol

## Availability of data and materials

Not applicable to this protocol

## Funding

This research is funded by the Clinton Health Access Initiative (CHAI) via funding to the Lancet Global Health Commission on Medical Oxygen.

CHAI Project ID: CHGEMCOVID14 CHAI Grant ID: UNITAIDCOVID6

Date: May 23rd, 2023

