## Supplementary Material for "The influence of skin pigmentation on pulse oximetry readings: a protocol for systematic review and meta-analysis"

**Search Strategy**

Ovid MEDLINE(R) ALL <1946 to September 28, 2023>

1. oximetry.mp. [mp=title, book title, abstract, original title, name of substance word, subject heading word, floating sub-heading word, keyword heading word, organism supplementary concept word, protocol supplementary concept word, rare disease supplementary concept word, unique identifier, synonyms, population supplementary concept word, anatomy supplementary concept word]	20493
2. exp Oximetry/	16739
3. (oximet* or oxymet*).ti,ab,kw.	18930
4. (SpO2 or %spo2 or sp o2).tw.	8070
5. 2 or 3 or 4	31921
6. (co-oximet* or co-oxymet* or h?emoximet*).ti,ab,kw.	855
7. (blood adj3 (analys* or measure*)).tw.	119958
8. (blood sampl* or gold standard or reference device* or reference instrument* or in-line oximet* or in vitro oximet* or arterial oxygen saturation or arterial oxyhemoglobin saturation or arterial oxyhaemoglobin saturation or arterial blood or arterial puncture or SaO2 or %SaO2 or sa o2).tw.	331459
9. 6 or 7 or 8	426217
10. Reproducibility of Results/	465554
11. Validation Study/	109229
12. Evaluation Studies as Topic/	122473
13. Bias/	26792
14. "Sensitivity and Specificity"/	369458
15. Hypoxia/di [Diagnosis]	2418
16. comparative study.pt.	1913099
17. (accura* or inaccura* or overestimat* or over-estimat* or underestimat* or under-estimat* or agreement or root-mean-square or root mean square or RMS or quadratic mean).tw.	1480376
18. (precision or evaluat* or predict* or reliab* or reproducib* or concordance or performance or bias or validat* or error* or erroneous or individual variability or (variability and (analysis or values)) or sensitivity or specificity or failure).tw.	9275801
19. (compar* adj3 (measure* or value*)).tw.	164286
20. (controlled desaturation or paired repeated measure* or method comparison or calibration stud*).ti,ab,kw.	3021
21. (paired readings or paired measurements or "difference of values" or "limits of agreement" or "limits of values" or confidence limits or regression or bland altman).ti,ab,kw.	1075980
22. 10 or 11 or 12 or 13 or 14 or 15 or 16 or 17 or 18 or 19 or 20 or 21	11573572
23. 5 and 9 and 22	4537
24. limit 23 to (english language and humans and yr="2022 -Current")	196

Embase Classic+Embase <1947 to 2023 Week 38>

1. exp Oximetry/	36304
2. (oximet* or oxymet*).ti,ab,kw.	27831
3. (SpO2 or %spo2 or sp o2).tw.	16827
4. 1 or 2 or 3	58489
5. (co-oximet* or co-oxymet* or h?emoximet*).ti,ab,kw.	1319
6. (blood adj3 (analys* or measure*)).tw.	180405
7. (blood sampl* or gold standard or reference device* or reference instrument* or in-line oximet* or in vitro oximet* or arterial oxygen saturation or arterial oxyhemoglobin saturation or arterial oxyhaemoglobin saturation or arterial blood or arterial puncture or SaO2 or %SaO2 or sa o2).tw.	516665
8. 5 or 6 or 7	658509
9. Reproducibility of Results/	210769
10. Validation Study/	106971
11. Evaluation Studies as Topic/	47038
12. Bias/	7858
13. "Sensitivity and Specificity"/	489707
14. Hypoxia/di [Diagnosis]	1236
15. comparative study.pt.	0
16. (accura* or inaccura* or overestimat* or over-estimat* or underestimat* or under-estimat* or agreement or root-mean-square or root mean square or RMS or quadratic mean).tw.	1899456
17. (precision or evaluat* or predict* or reliab* or reproducib* or concordance or performance or bias or validat* or error* or erroneous or individual variability or (variability and (analysis or values)) or sensitivity or specificity or failure).tw.	12508696
18. (compar* adj3 (measure* or value*)).tw.	229991
19. (controlled desaturation or paired repeated measure* or method comparison or calibration stud*).ti,ab,kw.	5228
20. (paired readings or paired measurements or "difference of values" or "limits of agreement" or "limits of values" or confidence limits or regression or bland altman).ti,ab,kw.	1537205
21. 9 or 10 or 11 or 12 or 13 or 14 or 15 or 16 or 17 or 18 or 19 or 20	13877475
22. 21 and 4 and 8	6652
23. limit 22 to (human and english language and yr="2022 -Current")	554

**Data extraction sheet for “Influence of skin tone on pulse oximetry readings”**

| Sl # | Item | Option | | |  |
| --- | --- | --- | --- | --- | --- |
| 1. 1 | Study ID (Reviewer’s initials) |  | | | Initials of reviewer |
| 2. 2 | Study Title |  | | | Text |
| 3. 3 | First Author Last Name |  | | | Text |
| 4. 4 | Last Author Last Name |  | | | Text |
| 5. 7 | Name of the Journal |  | | | Text |
| 6. 5 | Year of Publication |  | | | YYYY |
| 7. 6 | Type of publication | Original article | 1 |  |  |
|  |  | Review article | 2 |  |  |
|  |  | Case series | 3 |  |  |
|  |  | Letter to editor | 4 |  |  |
|  |  | Commentary | 5 |  |  |
|  |  | Other | 6 |  |  |
| 8. | Type of publication (other) |  | | | Text |
| 9. | Study start date |  | | | YYYY-MM |
| 10. | Study end date |  | | | YYYY-MM |
| 11. 8 | Type of Study | Cross Sectional | 1 |  |  |
|  |  | Case Control | 2 |  |  |
|  |  | Cohort | 3 |  |  |
|  |  | Individual RCT | 4 |  |  |
|  |  | Cluster RCT | 5 |  |  |
|  |  | Quasi-Experimental | 6 |  |  |
|  |  | Other | 7 |  |  |
| 12. | Type of study (other) |  | | | Text |
| 13. 9 | Study design | Prospective | 1 |  |  |
|  |  | Retrospective | 2 |  |  |
| 14. 13 | Study Settings-Region | Asia | 1 |  |  |
|  |  | Europe | 2 |  |  |
|  |  | Africa | 3 |  |  |
|  |  | South America | 4 |  |  |
|  |  | North America | 5 |  |  |
|  |  | Other | 6 |  |  |
| 15. | Study Settings-Region (other) |  | | | Text |
| 16. 22 | Country |  | | | Text |
| 17. 14 | Study Settings-Clinical | Inpatient | 1 |  |  |
|  |  | Outpatient | 2 |  |  |
|  |  | Laboratory setting | 3 |  |  |
|  |  | Emergency | 4 |  |  |
|  |  | Community | 5 |  |  |
|  |  | Other | 6 |  |  |
|  |  | Multiple | 7 |  |  |
|  |  | Not mentioned | 8 |  |  |
| 18. | Study Settings- Clinical (other) |  |  | | Text |
| 19. | Age range |  | | | Text |
| 20. 15 | Age group | 0-5 years | 1 |  |  |
|  |  | 6-18 year | 2 |  |  |
|  |  | 19-65 years | 3 |  |  |
|  |  | >65 years | 4 |  |  |
|  |  | Other range | 5 |  |  |
| 21. | Age group (other) |  | | | Text |
| 22. 16 | Average age of participants (Mean) |  | | | Numerical |
| 23. | Standard deviation of mean (Age) |  | | | Numerical |
| 24. | Average age of participants (Median) |  | | | Numerical |
| 25. | IQR of median (Age) |  | | | Text |
| 26. 19 | Number of participants |  | | | Numerical |
| 27. 17 | Male sex (Number) |  | | | Numerical |
| 28. 18 | Female sex (Number) |  | | | Numerical |
| 29. 20 | Skin pigmentation reported |  | Yes | No |  |
| 30. | Skin pigmentation Type | Light | 1 |  |  |
|  |  | Medium | 2 |  |  |
|  |  | Dark | 3 |  |  |
|  |  | Multiple | 4 |  |  |
| 31. 23 | Clinical condition of participants |  | | | Text |
| 32. | Type of pulse oximeter mentioned |  | Yes | No |  |
| 33. | Type of pulse oximeter | Fingertip | 1 |  |  |
|  |  | Handheld | 2 |  |  |
|  |  | Tabletop | 3 |  |  |
|  |  | Other | 4 |  |  |
|  |  | Multiple | 5 |  |  |
| 34. | Type of pulse oximeter (other) |  |  |  | Text |
| 35. | Model of pulse oximeter (details) |  |  |  | Text |
| 36. | Location of sensor (probe site) mentioned | Ear | 1 |  |  |
|  |  | Toe | 2 |  |  |
|  |  | Finger | 3 |  |  |
|  |  | Other | 4 |  |  |
|  |  | Not mentioned | 5 |  |  |
|  |  | Multiple | 6 |  |  |
| 37. | Location of sensor (other) |  |  |  | Text |
| 38. 27 | Method used for measuring SaO2 | Blood gas analyser | 1 |  |  |
|  |  | CO-oximeter model | 2 |  |  |
|  |  | Multiple | 3 |  |  |
|  |  | Other | 4 |  |  |
|  |  | Not mentioned | 5 |  |  |
| 39. | Method used for measuring SaO2 (other) |  |  |  | Text |
| 40. 28 | Blood source | Radial artery | 1 |  |  |
|  |  | Brachial artery | 2 |  |  |
|  |  | Multiple | 3 |  |  |
|  |  | Other | 4 |  |  |
|  |  | Not mentioned | 5 |  |  |
| 41. | Blood source (other) |  |  |  | Text |
| 42. | Skin pigmentation definition reported | Yes | 1 |  |  |
|  |  | No | 2 |  |  |
| 43. | Skin pigmentation definition used | Subjective scale | 1 |  |  |
|  |  | Fitzpatrick phototype | 2 |  |  |
|  |  | Munsell scale | 3 |  |  |
|  |  | Portable EEL reflectance spectrometer | 4 |  |  |
|  |  | Ancestry | 5 |  |  |
|  |  | Multiple | 6 |  |  |
|  |  | Other | 7 |  |  |
| 44. | Skin pigmentation definition used (other) |  |  |  | Text |

| 45. | Accuracy reported | Yes | 1 |  |  |
| --- | --- | --- | --- | --- | --- |
|  |  | No | 2 |  |  |
| 46. | Accuracy |  | | | Numerical |
| 47. | Precision reported | Yes | 1 |  |  |
|  |  | No | 2 |  |  |
| 48. | Precision |  | | | Numerical |
| 49. | Agreement reported | Yes | 1 |  |  |
|  |  | No | 2 |  |  |
| 50. | Agreement |  | | | Numerical |

| 51. | Bias reported | Yes | 1 |
| --- | --- | --- | --- |
|  |  | No | 2 |

| 52. | Bias mean difference |  | | | Numerical |
| --- | --- | --- | --- | --- | --- |
| 53. | Bias denominator |  | | | Numerical |
| 54. | Bias SD (Standard deviation) |  | | | Numerical |
| 55. | Bias Upper limit CI (Confidence Interval) |  | | | Numerical |
| 56. | Bias Lower limit CI (Confidence Interval) |  | | | Numerical |
| 57. | Bias Direction | SpO2- SaO2 | 1 |  |  |
|  |  | SaO2- SpO2 | 2 |  |  |
| 58. | Bias average SpO2 |  | | | Numerical |
| 59. | Bias average SaO2 |  | | | Numerical |

| 60. | Repeated measures design (more than one SpO2-SaO2 data pair collected per person) | Yes | 1 |  |  |
| --- | --- | --- | --- | --- | --- |
|  |  | No | 2 |  |  |
| **Disaggregated data** | | | | | |
| 61. 31 | **Disaggregated bias reported for age** | Yes | 1 |  |  |
|  |  | No | 2 |  |  |
| 62. 31 | **Bias reported for 0- 5 years** | Yes | 1 |  |  |
|  |  | No | 2 |  |  |
| 63. | Bias mean difference |  | | | Numerical |
| 64. | Bias denominator |  | | | Numerical |
| 65. | Bias SD (Standard deviation) |  | | | Numerical |
| 66. | Bias Upper limit CI (Confidence Interval) |  | | | Numerical |
| 67. | Bias Lower limit CI (Confidence Interval) |  | | | Numerical |

| 68. | Bias Direction | SpO2- SaO2 | 1 |
| --- | --- | --- | --- |
|  |  | SaO2- SpO2 | 2 |

| 69. | Bias average SpO2 |  | Numerical |
| --- | --- | --- | --- |
| 70. | Bias average SaO2 |  | Numerical |

| 71. 31 | **Bias reported for 6-18 years** | | Yes | | 1 | | |  | |  |
| --- | --- | --- | --- | --- | --- | --- | --- | --- | --- | --- |
|  |  |  | No | | 2 | | |  | |  |
| 72. | Bias mean difference | |  | | | | | | | Numerical |
| 73. | Bias denominator | |  | | | | | | | Numerical |
| 74. | Bias SD (Standard deviation) | |  | | | | | | | Numerical |
| 75. | Bias Upper limit CI (Confidence Interval) | |  | | | | | | | Numerical |
| 76. | Bias Lower limit CI (Confidence Interval) | |  | | | | | | | Numerical |
| 77. | Bias Direction | | SpO2- SaO2 | 1 | | | |  | |  |
|  |  |  | SaO2- SpO2 | 2 | | | |  | |  |
| 78. | Bias average SpO2 | |  | | | | | | | Numerical |
| 79. | Bias average SaO2 | |  | | | | | | | Numerical |
| 80. | **Bias reported for 19-65 years** | Yes | | | | | 1 | |  |  |
|  |  | No | | | | | 2 | |  |  |
| 81. | Bias mean difference |  | | | | | | | | Numerical |
| 82. | Bias denominator |  | | | | | | | | Numerical |
| 83. | Bias SD (Standard deviation) |  | | | | | | | | Numerical |
| 84. | Bias Upper limit CI (Confidence Interval) |  | | | | | | | | Numerical |
| 85. | Bias Lower limit CI (Confidence Interval) |  | | | | | | | | Numerical |
| 86. | Bias Direction | SpO2- SaO2 | | | | 1 | | |  |  |
|  |  | SaO2- SpO2 | | | | 2 | | |  |  |
| 87. | Bias average SpO2 |  | | | | | | | | Numerical |
| 88. | Bias average SaO2 |  | | | | | | | | Numerical |
| 89. | **Bias reported for >65 years** | Yes | | | | | 1 | |  |  |
|  |  | No | | | | | 2 | |  |  |
| 90. | Bias mean difference |  | | | | | | | | Numerical |
| 91. | Bias denominator |  | | | | | | | | Numerical |
| 92. | Bias SD (Standard deviation) |  | | | | | | | | Numerical |
| 93. | Bias Upper limit CI (Confidence Interval) |  | | | | | | | | Numerical |
| 94. | Bias Lower limit CI (Confidence Interval) |  | | | | | | | | Numerical |
| 95. | Bias Direction | SpO2- SaO2 | | | | 1 | | |  |  |
|  |  | SaO2- SpO2 | | | | 2 | | |  |  |
| 96. | Bias average SpO2 |  | | | | | | | | Numerical |
| 97. | Bias average SaO2 |  | | | | | | | | Numerical |
| 98. | Bias reported for other age categories | Yes | | | | | 1 | |  |  |
|  |  | No | | | | | 2 | |  |  |
| 99. | Bias mean difference |  | | | | | | | | Numerical |
| 100. | Bias denominator |  | | | | | | | | Numerical |
| 101. | Bias SD (Standard deviation) |  | | | | | | | | Numerical |
| 102. | Bias Upper limit CI (Confidence Interval) |  | | | | | | | | Numerical |
| 103. | Bias Lower limit CI (Confidence Interval) |  | | | | | | | | Numerical |
| 104. | Bias Direction | SpO2- SaO2 | | | | 1 | | |  |  |
|  |  | SaO2- SpO2 | | | | 2 | | |  |  |
| 105. | Bias average SpO2 |  | | | | | | | | Numerical |
| 106. | Bias average SaO2 |  | | | | | | | | Numerical |
| 107. 3 | **Disaggregated bias reported for sex** | Yes | | | | | 1 | |  |  |
|  |  | No | | | | | 2 | |  |  |
| 108. 3 | **Bias reported for male** | Yes | | | | | 1 | |  |  |
|  |  | No | | | | | 2 | |  |  |
| 109. | Bias mean difference |  | | | | | | | | Numerical |
| 110. | Bias denominator |  | | | | | | | | Numerical |
| 111. | Bias SD (Standard deviation) |  | | | | | | | | Numerical |
| 112. | Bias Upper limit CI (Confidence Interval) |  | | | | | | | | Numerical |
| 113. | Bias Lower limit CI (Confidence Interval) |  | | | | | | | | Numerical |
| 114. | Bias Direction | SpO2- SaO2 | | | | 1 | | |  |  |
|  |  | SaO2- SpO2 | | | | 2 | | |  |  |
| 115. | Bias average SpO2 |  | | | | | | | | Numerical |
| 116. | Bias average SaO2 |  | | | | | | | | Numerical |
| 117. 3 | **Bias reported for female** | Yes | | | | | 1 | |  |  |
|  |  | No | | | | | 2 | |  |  |
| 118. | Bias mean difference |  | | | | | | | | Numerical |
| 119. | Bias denominator |  | | | | | | | | Numerical |
| 120. | Bias SD (Standard deviation) |  | | | | | | | | Numerical |
| 121. | Bias Upper limit CI (Confidence Interval) |  | | | | | | | | Numerical |
| 122. | Bias Lower limit CI (Confidence Interval) |  | | | | | | | | Numerical |
| 123. | Bias Direction | SpO2- SaO2 | | | | 1 | | |  |  |
|  |  | SaO2- SpO2 | | | | 2 | | |  |  |
| 124. | Bias average SpO2 |  | | | | | | | | Numerical |
| 125. | Bias average SaO2 |  | | | | | | | | Numerical |
| 126. 3 | **Disaggregated bias reported for skin pigmentation** | Yes | | | | | 1 | |  |  |
|  |  | No | | | | | 2 | |  |  |
| 127. 3 | **Bias reported for dark skin** | Yes | | | | | 1 | |  |  |
|  |  | No | | | | | 2 | |  |  |
| 128. | Bias mean difference |  | | | | | | | | Numerical |
| 129. | Bias denominator |  | | | | | | | | Numerical |
| 130. | Bias SD (Standard deviation) |  | | | | | | | | Numerical |
| 131. | Bias Upper limit CI (Confidence Interval) |  | | | | | | | | Numerical |
| 132. | Bias Lower limit CI (Confidence Interval) |  | | | | | | | | Numerical |
| 133. | Bias Direction | SpO2- SaO2 | | | | 1 | | |  |  |
|  |  | SaO2- SpO2 | | | | 2 | | |  |  |
| 134. | Bias average SpO2 |  | | | | | | | | Numerical |
| 135. | Bias average SaO2 |  | | | | | | | | Numerical |
| 136. 3 | **Bias reported for medium skin** | Yes | | | | | 1 | |  |  |
|  |  | No | | | | | 2 | |  |  |
| 137. | Bias mean difference |  | | | | | | | | Numerical |
| 138. | Bias denominator |  | | | | | | | | Numerical |
| 139. | Bias SD (Standard deviation) |  | | | | | | | | Numerical |
| 140. | Bias Upper limit CI (Confidence Interval) |  | | | | | | | | Numerical |
| 141. | Bias Lower limit CI (Confidence Interval) |  | | | | | | | | Numerical |
| 142. | Bias Direction | SpO2- SaO2 | | | | 1 | | |  |  |
|  |  | SaO2- SpO2 | | | | 2 | | |  |  |
| 143. | Bias average SpO2 |  | | | | | | | | Numerical |
| 144. | Bias average SaO2 |  | | | | | | | | Numerical |
| 145. 3 | **Bias reported for light skin** | Yes | | | | | 1 | |  |  |
|  |  | No | | | | | 2 | |  |  |
| 146. | Bias mean difference |  | | | | | | | | Numerical |
| 147. | Bias denominator |  | | | | | | | | Numerical |
| 148. | Bias SD (Standard deviation) |  | | | | | | | | Numerical |
| 149. | Bias Upper limit CI (Confidence Interval) |  | | | | | | | | Numerical |
| 150. | Bias Lower limit CI (Confidence Interval) |  | | | | | | | | Numerical |
| 151. | Bias Direction | SpO2- SaO2 | | | | 1 | | |  |  |
|  |  | SaO2- SpO2 | | | | 2 | | |  |  |
| 152. | Bias average SpO2 |  | | | | | | | | Numerical |
| 153. | Bias average SaO2 |  | | | | | | | | Numerical |
| 154. | **Disaggregated bias reported for ethnicity** | Yes | | | | | 1 | |  |  |
|  |  | No | | | | | 2 | |  |  |
| 155. 3 | **Bias reported for Black/African** | Yes | | | | | 1 | |  |  |
|  |  | No | | | | | 2 | |  |  |
| 156. | Bias mean difference |  | | | | | | | | Numerical |
| 157. | Bias denominator |  | | | | | | | | Numerical |
| 158. | Bias SD (Standard deviation) |  | | | | | | | | Numerical |
| 159. | Bias Upper limit CI (Confidence Interval) |  | | | | | | | | Numerical |
| 160. | Bias Lower limit CI (Confidence Interval) |  | | | | | | | | Numerical |
| 161. | Bias Direction | SpO2- SaO2 | | | | 1 | | |  |  |
|  |  | SaO2- SpO2 | | | | 2 | | |  |  |
| 162. | Bias average SpO2 |  | | | | | | | | Numerical |
| 163. | Bias average SaO2 |  | | | | | | | | Numerical |
| 164. 3 | **Bias reported for Asian, Hispanic, mixed** | Yes | | | | | 1 | |  |  |
|  |  | No | | | | | 2 | |  |  |
| 165. | Bias mean difference |  | | | | | | | | Numerical |
| 166. | Bias denominator |  | | | | | | | | Numerical |
| 167. | Bias SD (Standard deviation) |  | | | | | | | | Numerical |
| 168. | Bias Upper limit CI (Confidence Interval) |  | | | | | | | | Numerical |
| 169. | Bias Lower limit CI (Confidence Interval) |  | | | | | | | | Numerical |
| 170. | Bias Direction | SpO2- SaO2 | | | | 1 | | |  |  |
|  |  | SaO2- SpO2 | | | | 2 | | |  |  |
| 171. | Bias average SpO2 |  | | | | | | | | Numerical |
| 172. | Bias average SaO2 |  | | | | | | | | Numerical |
| 173. 3 | **Bias reported for White/ Caucasian** | Yes | | | | | 1 | |  |  |
|  |  | No | | | | | 2 | |  |  |
| 174. | Bias mean difference |  | | | | | | | | Numerical |
| 175. | Bias denominator |  | | | | | | | | Numerical |
| 176. | Bias SD (Standard deviation) |  | | | | | | | | Numerical |
| 177. | Bias Upper limit CI (Confidence Interval) |  | | | | | | | | Numerical |
| 178. | Bias Lower limit CI (Confidence Interval) |  | | | | | | | | Numerical |
| 179. | Bias Direction | SpO2- SaO2 | | | | 1 | | |  |  |
|  |  | SaO2- SpO2 | | | | 2 | | |  |  |
| 180. | Bias average SpO2 |  | | | | | | | | Numerical |
| 181. | Bias average SaO2 |  | | | | | | | | Numerical |
| 182. 3 | **Bias reported for other factors** | Yes | | | | | 1 | |  |  |
|  |  | No | | | | | 2 | |  |  |
| 183. | Bias mean difference |  | | | | | | | | Numerical |
| 184. | Bias denominator |  | | | | | | | | Numerical |
| 185. | Bias SD (Standard deviation) |  | | | | | | | | Numerical |
| 186. | Bias Upper limit CI (Confidence Interval) |  | | | | | | | | Numerical |
| 187. | Bias Lower limit CI (Confidence Interval) |  | | | | | | | | Numerical |
| 188. | Bias Direction | SpO2- SaO2 | | | | 1 | | |  |  |
|  |  | SaO2- SpO2 | | | | 2 | | |  |  |
| 189. | Bias average SpO2 |  | | | | | | | | Numerical |
| 190. | Bias average SaO2 |  | | | | | | | | Numerical |
| 191. | Additional comments |  | | | | | | | | Text |
| 192. | SpO2 reported? |  | | | | | | | |  |
| 193. A | SaO2 reported? |  | | | | | | | |  |
| 194. | Details of SaO2 machine and procedure |  | | | | | | | |  |
